# Analysis of Image-Guided Superficial Radiation Therapy (IGSRT) on the Treatment of Early Stage Non-Melanoma Skin Cancer (NMSC) in the Outpatient Dermatology Setting

**DOI:** 10.1101/2022.09.14.22279951

**Authors:** Alison Tran, Mairead Moloney, Peter Kaczmarksi, Songzhu Zheng, Alpesh Desai, Tejas Desai, Lio Yu

## Abstract

Interest in image-guidance superficial radiation therapy (IGSRT) for the treatment of early-stage Non-Melanoma Skin Cancer (NMSC) has resurfaced given its low complication rates, superior cosmesis and local control and cure rates. Additionally, it has been recommended by the American Academy of Dermatology (AAD) for early-stage NMSC in patients who are considered poor surgical candidates.

**Methods:** 1899 NMSC lesions were treated with energies ranging from 50 to 100 kilovoltage (kV), for a mean of 20.2 fractions, and treatment dose of 5364.4 centigray (cGy). Lesions were treated for a mean of 7.5 weeks and followed for 65.5 weeks. SAS studio was used to conduct Kaplan-Meier analysis to calculate local control rates and account for differences in follow-up intervals. A log-rank test was used to calculate statistical differences between histologies.

**Results:** Absolute lesion control was achieved in 99.7% of the patients after an average of 7.5 weeks of treatment, with a stable control rate of 99.6% when the follow-up duration was over 12 months.

**Conclusion:** IGSRT has a high safety profile, can achieve superior cosmesis and should be considered first-line for treating early-stage NMSC tumors as cure rates have been shown to be effective in all NMSC on early follow-up.

## 1.0 Introduction

Skin cancer is the most common form of cancer in the United States, with more than 9,500 people diagnosed daily [1]. The incidence of both melanoma and non-melanoma skin cancer (NMSC) has been steadily rising. Between 1994-2014, the diagnosis and treatment of NMSC in the United States increased by 77% [2]. NMSC is the most frequently diagnosed cancer, with an 18-20 times higher incidence than melanoma [3, 4, 5]. The most recent estimate in 2012 revealed more than 5.4 million cases of NMSC treated in over 3.3 million patients [4]. The cost of skin cancer treatment in the United States is estimated at $8.1 billion annually [6], of which approximately 4.3 million patients are treated for basal cell carcinoma (BCC) and squamous cell carcinoma (SCC) at $4.8 billion, while melanoma treatment costs $3.3 billion [3, 6]. However, these estimates do not consider those who have no access to treatment or are uninsured.

Skin cancers may arise from any host cell of the skin. However, BCC and SCC are keratinocyte carcinomas that account for 99% of NMSC [3, 4]. Overall, BCC is the most common form of skin cancer, followed by SCC [1]. However, SCC is the most common skin malignancy among African Americans and Asian Indians [6, 7]. BCC contributes to 65%-75% of skin cancers in whites and 20%-30% in people of color [6]. An estimated 5.03 to 5.23 million BCC lesions and between 200,000 to 400,000 SCC lesions are diagnosed yearly in the United States [4]. Central cancer registries do not generally collect data on basal cell and squamous cell carcinomas [7]. As such, the reported figures may be an underestimate.

The overall prognosis for both BCC and SCC is good, especially when detected at early stages [3]. BCC minimally contributes to the NMSC mortality rate (MR) at 0.02 per 10,000 [3, 8, 9]. Meanwhile, SCC shows a variable metastatic rate of 0.1–9.9%, accounting for about 75% of NMSC-related deaths [3, 8, 9]. Despite the high occurrence of NMSC, they are considered nonfatal and curable due to their slow growth, low recurrence, and rare metastasis [10,11]. Nonetheless, NMSC should be treated to prevent growth, invasion, and potential mortality. The latest data suggests that greater than 15,000 people die of SCC in the U.S. yearly, which is twice that of melanoma-related deaths [12] with more than 5,400 people worldwide dying of NMSC every month [13]. This translates to about 65,000 NMSC related deaths worldwide annually.

Major risk factors for the development of NMSC include increased exposure to ultraviolet (UV) radiation (hence the greatest incidence in sun-exposed areas, i.e., head and neck), as approximately 90% of NMSC are associated with UV exposure [1]. Other risk factors include older age, fair skin/hereditary risk factors, and improved surveillance, which contribute to earlier recognition [14]. In addition, genetic polymorphisms also modulate susceptibility to skin cancer [6, 15]. Chronic scarring and areas of chronic inflammation may also pose a risk factor for developing SCC in darkly pigmented individuals [6]. Moreover, SCCs have a greater tendency to occur in non-exposed sites with a higher potential for metastasis in Asians [6, 16].

Ethnic minorities, elderly, the less educated, uninsured and those of low socioeconomic status have poorer melanoma and NMSC outcomes [17]. Advanced stage at presentation, atypical distribution of malignant skin lesions and socioeconomic factors, e.g., lack of adequate insurance coverage and/or transportation prevent timely diagnosis and early treatment [18]. Though quality of care has reportedly improved throughout the years, access to care and health disparities have not [17]. Inadequate access to dermatologic care may be explained by the current dermatology workforce shortage coupled with the increased patient load [17]. Further, most counties with African, Hispanic and Native American majorities have no dermatologists [19, 20].

## 2.0 Treatment Modalities

Early stage NSMC treatments include Photodynamic Therapy (PDT), cryotherapy, topical medications such as imiquimod 5% and diclofenac sodium 3% [4], laser, electrodessication and curettage, and radiotherapies such as superficial radiation therapy (SRT), image-guided superficial radiation therapy (IGSRT), external beam radiation therapy (XRT) which include electron-beam radiation and isotope-based and electronic brachytherapy. Since the most frequently affected areas include the head and neck, it is essential to employ treatments that have high cure rates and simultaneously engender superior aesthetic results. While the standard of care is surgical excision, preferentially Mohs Micrographic Surgery (MMS), due to high cure rates and ability to achieve tissue conservation, not all patients are surgical candidates. In these situations, it is appealing to choose a non-surgical modality that has superior aesthetic results and high cure rates.

### 2.1 Superficial Radiation Therapy (SRT)

Superficial radiation therapy (SRT) has been used for decades to treat NMSC. It is a form of external radiotherapy that uses low energy, low penetration, kilovoltage (kV) photons between 50 to 150 kilovoltage peak (kVp) [21], which preferentially targets tumors of the skin while sparing deeper structures [21, 22] beyond the dermis. Recent advancements in radiation technology, i.e., better tumor depth coverage and use of image guidance, have improved local control and cure rates. Its low complication rates, e.g., no pain or scarring combined with superior cosmesis, make it an attractive alternative to surgical intervention. Hence, it has been recommended by the American Academy of Dermatology (AAD) [22] and studies published in the Journal of the American Academy of Dermatology (JAAD) [23] for the treatment of NMSC in patients who are poor surgical candidates and in other studies as a primary option in appropriate patients [24, 25].

### 2.2 Image-guided superficial radiation therapy (IGSRT)

IGSRT combines ultrasound technology with SRT delivery. The high resolution ultrasound (HRUS) is designed to detect dermatologic structures using frequencies of 22 MHz, which enable visualization of skin structures, lesions and depth as well as the lateral configuration of the tumor. The tumor depth is used to correlate with the percentage depth dose (PDD), which determines the selection of energy (50, 70, or 100 kV) delivered, and adjustments can be made during the treatment. The most recent and largest modern study (2917 cases of early-stage SCCIS, T1 and T2 NMSC) conducted by Yu and colleagues in 2021 demonstrated that IGSRT had an absolute local control rate of 99.3%, which was stably unchanged at the follow-up intervals of greater than one year to a max of 4 years [24].

## 3.0 Methods

A retrospective chart review of 1243 patients with a total of 1899 lesions from an outpatient dermatology practice in Dallas, Texas, was analyzed after the study protocol was reviewed and determined to be exempt from IRB approval by an IRB committee (WIRB-Copernicus Group) under 45 CFR 46.104 (d)(4). The information obtained was recorded by the investigator in such a manner that the identity of the human subject could not be readily ascertained directly or through identifiers linked to the subjects, the investigator does not contact the subjects, and the investigator will not re-identify subjects. Any health information used in this study has been de-identified. This study was performed in compliance with the pertinent sections of the Helsinki Declaration and its amendments. All methods were carried out in accordance with relevant guidelines and regulations.

### 3.1 Inclusion Criteria and Patient Demographics

Patients with varying Fitzpatrick skin types and NMSC, i.e., BCC, SCC and SCCIS who received twenty or more treatments were included in this study. Lesions that were not considered were keloids, non-keratinocytic tumors, and tumors of stage III or greater.

A majority (58.7%) of the patients identified as White, unspecified (40.9%), followed by American Indian/Alaska Native and Asian (0.2% and 0.1%, respectively). The majority of patients identified as Non-Hispanic or non-Latino consisting 54.6% of the study population, with 44.3% unspecified, while 1.0% of patients identified as Hispanic or Latino. Additionally, male patients contributed to 62.6% of the sample, while female patients comprised 36.7% with 0.7% unspecified. Lastly, the mean age of the sample was 73.2 years (SD ± 10.99 years), with 33.1 years being the youngest and 98.2 years being the oldest [Table 1].

**Table 1.**
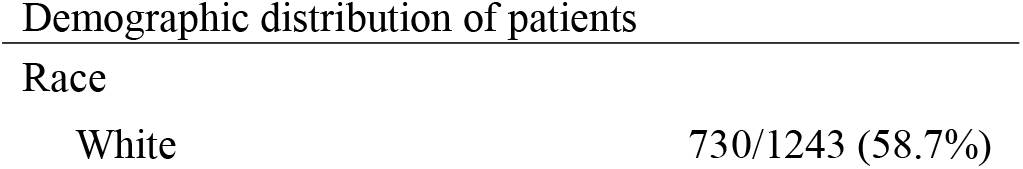
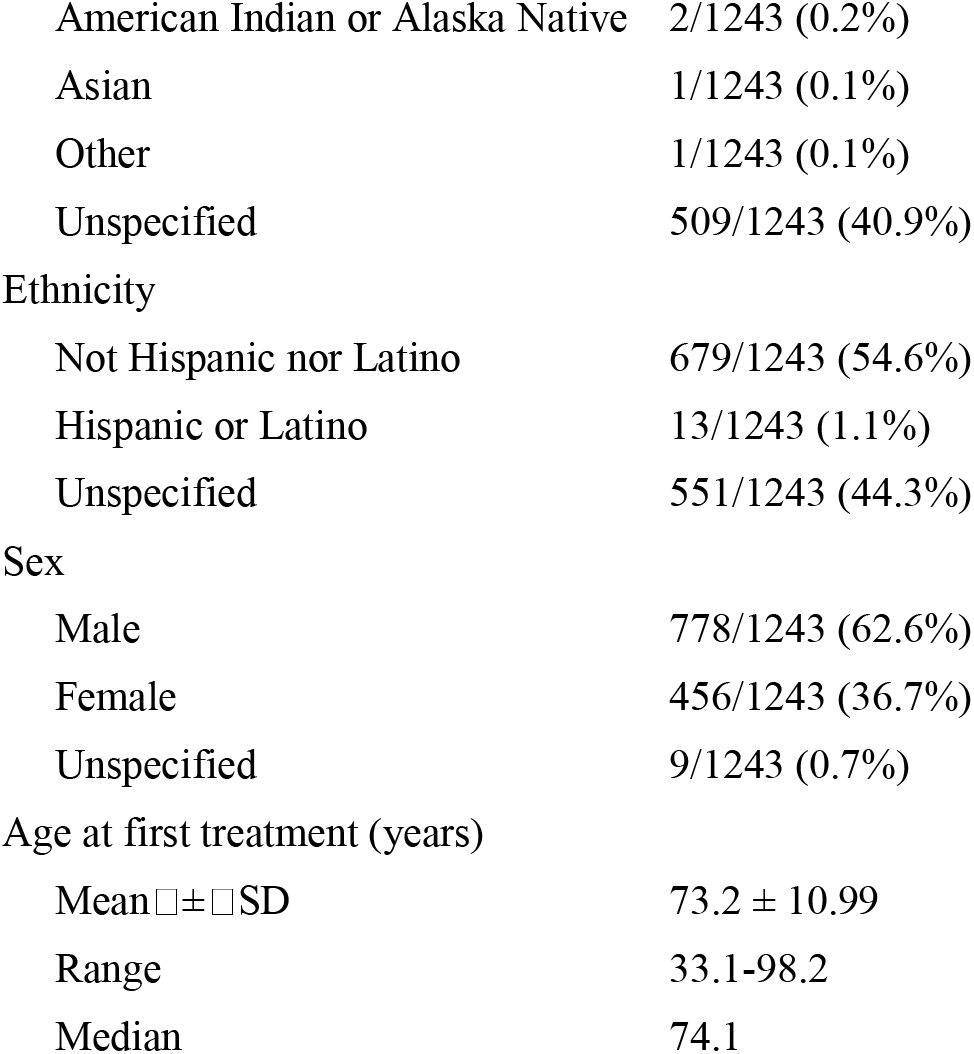
Demographic distribution of patients

| Race |  |
| --- | --- |
| White | 730/1243 (58.7%) |
| American Indian or Alaska Native | 2/1243 (0.2%) |
| Asian | 1/1243 (0.1%) |
| Other | 1/1243 (0.1%) |
| Unspecified | 509/1243 (40.9%) |
| Ethnicity |  |
| Not Hispanic nor Latino | 679/1243 (54.6%) |
| Hispanic or Latino | 13/1243 (1.1%) |
| Unspecified | 551/1243 (44.3%) |
| Sex |  |
| Male | 778/1243 (62.6%) |
| Female | 456/1243 (36.7%) |
| Unspecified | 9/1243 (0.7%) |
| Age at first treatment (years) |  |
| Mean $\bar{x} \pm \text{SD}$ | 73.2 ± 10.99 |
| Range | 33.1-98.2 |
| Median | 74.1 |

### 3.2 Treatment

Board-Certified radiation therapists administered IGSRT technology to treat lesions with energies ranging from 50, 70 or 100 kilovoltage (kV), which was delivered 2-4 times weekly. The mean total number of fractions was 20.2 (SD ± 0.90), ranging from 20 to 30. The mean total treatment dose was 5364.4 centigray (cGy) (SD ± 241.60), ranging from 4453.4 to 6703.2 cGy. The majority of these lesions were treated for 7.5 weeks and followed for a mean of 65.5 weeks (□ SD± □66.70) [Table 2]. The duration of follow-up was calculated as the date of last follow-up minus the last treatment date plus one day.

**Table 2.**
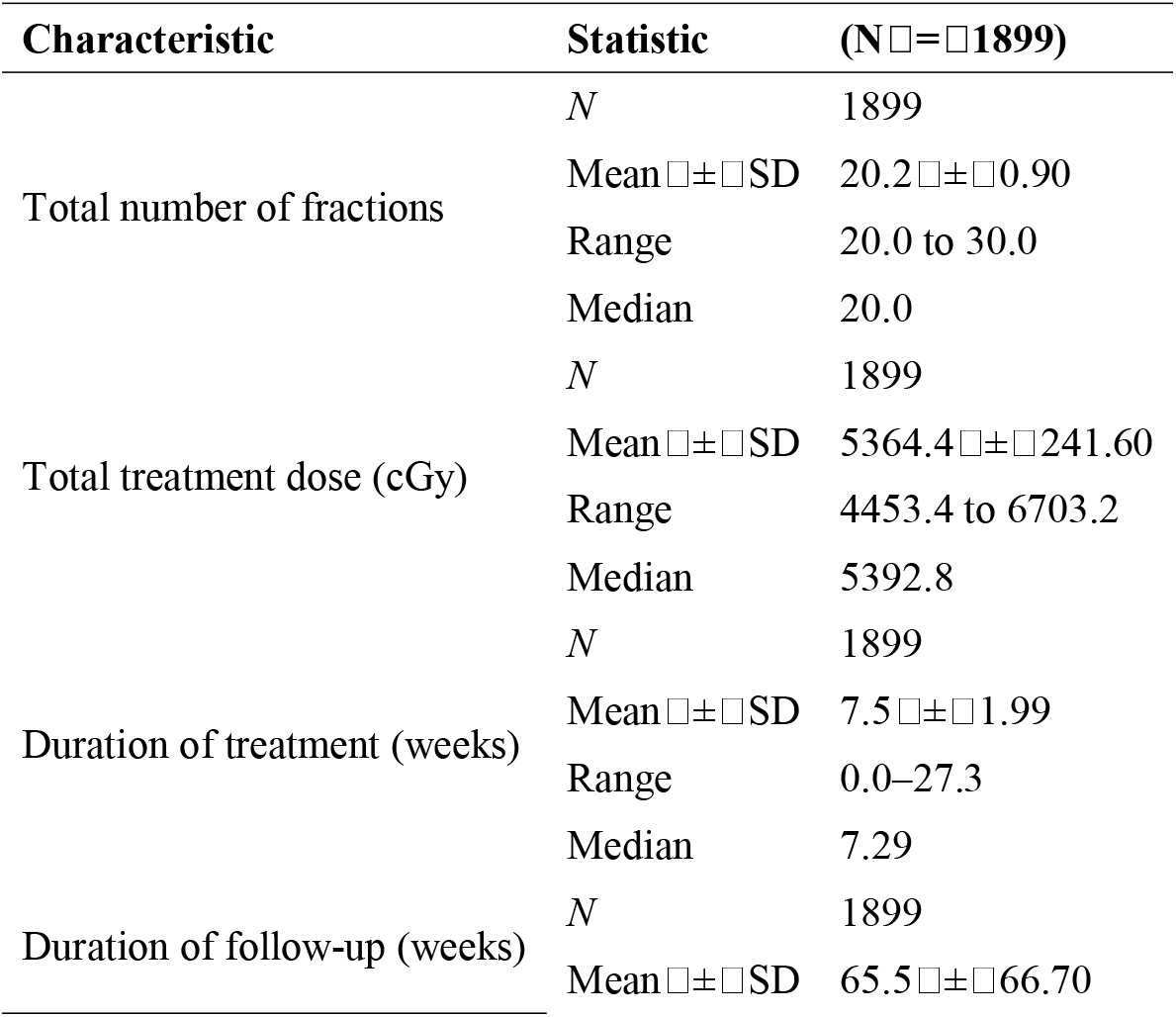
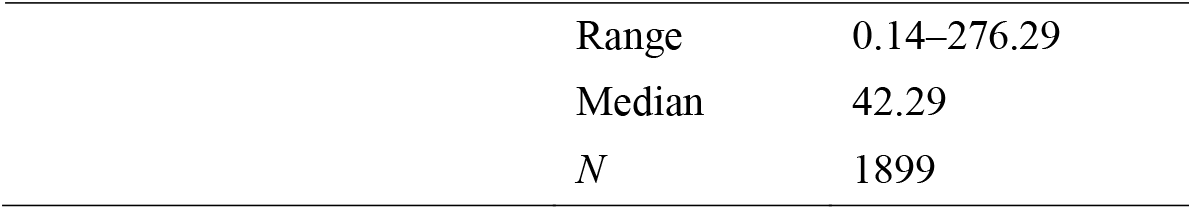
Total number of treatment fractions, dose, duration and follow-up interval

Energy selection and dose adjustments were contingent upon tumor characteristics seen clinically and on ultrasound (histology and depth). One hundred seventy-six lesions (9.9% [176/1779]) were treated with a combination of two or more energies. Lesion treatment by energy is summarized in the following [Table 3]. The Radiation Treatment Oncology Group (RTOG) toxicity scoring system was used to grade acute toxicities after every 5 fractions and the highest RTOG score was recorded. These on treatment evaluations occurred throughout the treatment course.

**Table 3.** NMSC lesion treatment by energy

| Energy (kV) | BCC only | SCC only | SCCIS only | Combined BCC and SCC | Combined BCC and SCCIS | Number of lesions<sup>a</sup> (N=1779) |
| --- | --- | --- | --- | --- | --- | --- |
| 50 | 200 | 70 | 79 | 0 | 0 | 349 |
| 70 | 456 | 236 | 257 | 0 | 1 | 950 |
| 100 | 175 | 73 | 55 | 1 | 0 | 304 |
| Mixed | 94 | 50 | 32 | 0 | 0 | 176 |

<sup>a</sup>116 lesions with no energy data and 4 lesions with no histology data.

Follow-up occurred at 2-12 week intervals with a mean follow-up of 65.5 weeks after treatment completion until No Evidence of Disease (NED) was achieved or failure/recurrence. Safety checks on the SRT machine were performed by the medical physicist regularly.

### 3.3 Statistical Analysis

Missing data were excluded from final analysis. SAS studio was used to conduct Kaplan-Meier analysis to calculate local control rates and account for differences in follow-up intervals between patients. A log-rank test was used to calculate statistical differences between histologies with a p-value of <0.05 considered statistically significant.

## 4.0 Results

In this current study, a total of 1899 NMSC lesions in 1243 patients were treated with IGSRT from 2016 to 2022. The cohort consisted of 981 BCC, 467 SCC, and 444 SCCIS lesions (certain lesions had combinations of two or more of these histologies with five lesions of unspecified histology. Among 1243 patients, 99.7% (1239/1243) were alive as of May 2022. All deaths were deemed unrelated to the treatment of NMSC by IGSRT.

Table 4 shows the most common sites of the treated lesions were head and neck as a group, extremities, followed by the head and neck subgroups cheek and nose. Table 5 demonstrates that BCC (51.7%) was more common than SCC (24.6%) and SCCIS (23.4%). The mean diameter of measured lesions was 1.3 cm (SD ± 0.69), ranging from 0.0 to 3.9 cm. The mean diameter was 1.3 mm (SD ± 0.70) for BCC, 1.4 mm (SD ± 0.66) for SCC, and 1.3mm (SD ± 0.68) for SCCIS. Of the 981 BCC lesions, the measured diameter ranging from 0 to <2 cm (T1) was detected in approximately 78.1% of lesions and 19.3% of lesions were observed to have diameters ranging from 2 to <4 cm (T2). Of the 467 SCC lesions, the measured diameter ranging from 0 to <2 cm was observed in 73.2% of lesions, and diameters ranging from 2 to <4 cm were seen in 24.4% of lesions. 76.1% of the 444 total SCCIS lesions (Tis) were observed to have a diameter ranging from 0 to <2 cm and 20.5% of lesions were found to have diameters ranging from 2 to <4 cm. SCCIS lesions greater than or equal to 4 cm were still considered Tis by AJCC staging manual 8^th^ edition [26] [Table 6].

**Table 4.**
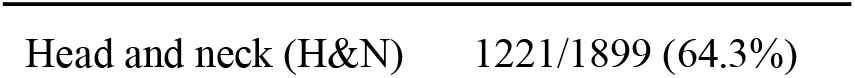
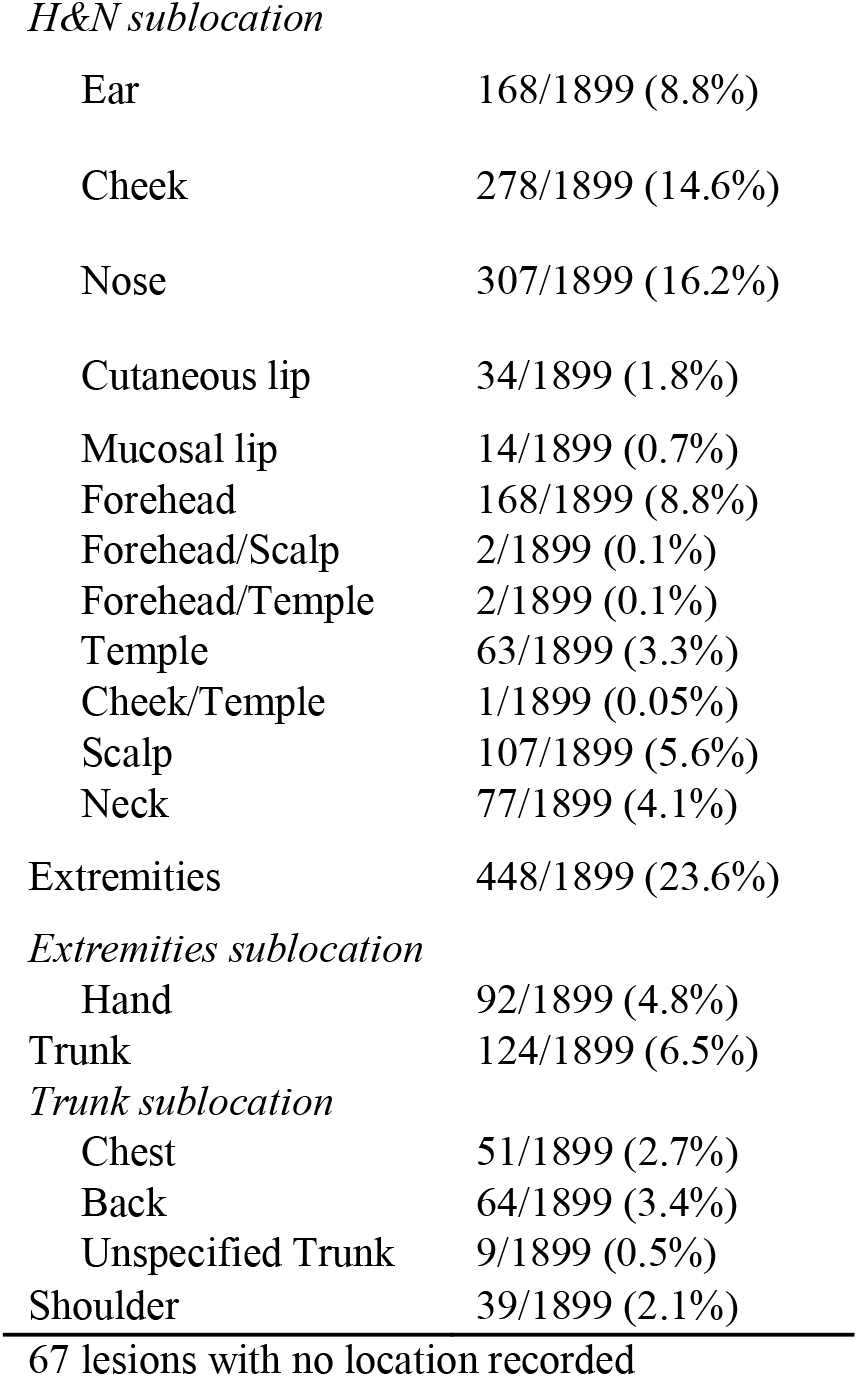
Anatomic distribution of NMSC lesions

|  |  |
| --- | --- |
| Head and neck (H&N) | 1221/1899 (64.3%) |
| *H&N sublocation* |  |
| Ear | 168/1899 (8.8%) |
| Cheek | 278/1899 (14.6%) |
| Nose | 307/1899 (16.2%) |
| Cutaneous lip | 34/1899 (1.8%) |
| Mucosal lip | 14/1899 (0.7%) |
| Forehead | 168/1899 (8.8%) |
| Forehead/Scalp | 2/1899 (0.1%) |
| Forehead/Temple | 2/1899 (0.1%) |
| Temple | 63/1899 (3.3%) |
| Cheek/Temple | 1/1899 (0.05%) |
| Scalp | 107/1899 (5.6%) |
| Neck | 77/1899 (4.1%) |
| Extremities | 448/1899 (23.6%) |
| *Extremities sublocation* |  |
| Hand | 92/1899 (4.8%) |
| Trunk | 124/1899 (6.5%) |
| *Trunk sublocation* |  |
| Chest | 51/1899 (2.7%) |
| Back | 64/1899 (3.4%) |
| Unspecified Trunk | 9/1899 (0.5%) |
| Shoulder | 39/1899 (2.1%) |

67 lesions with no location recorded

**Table 5.**
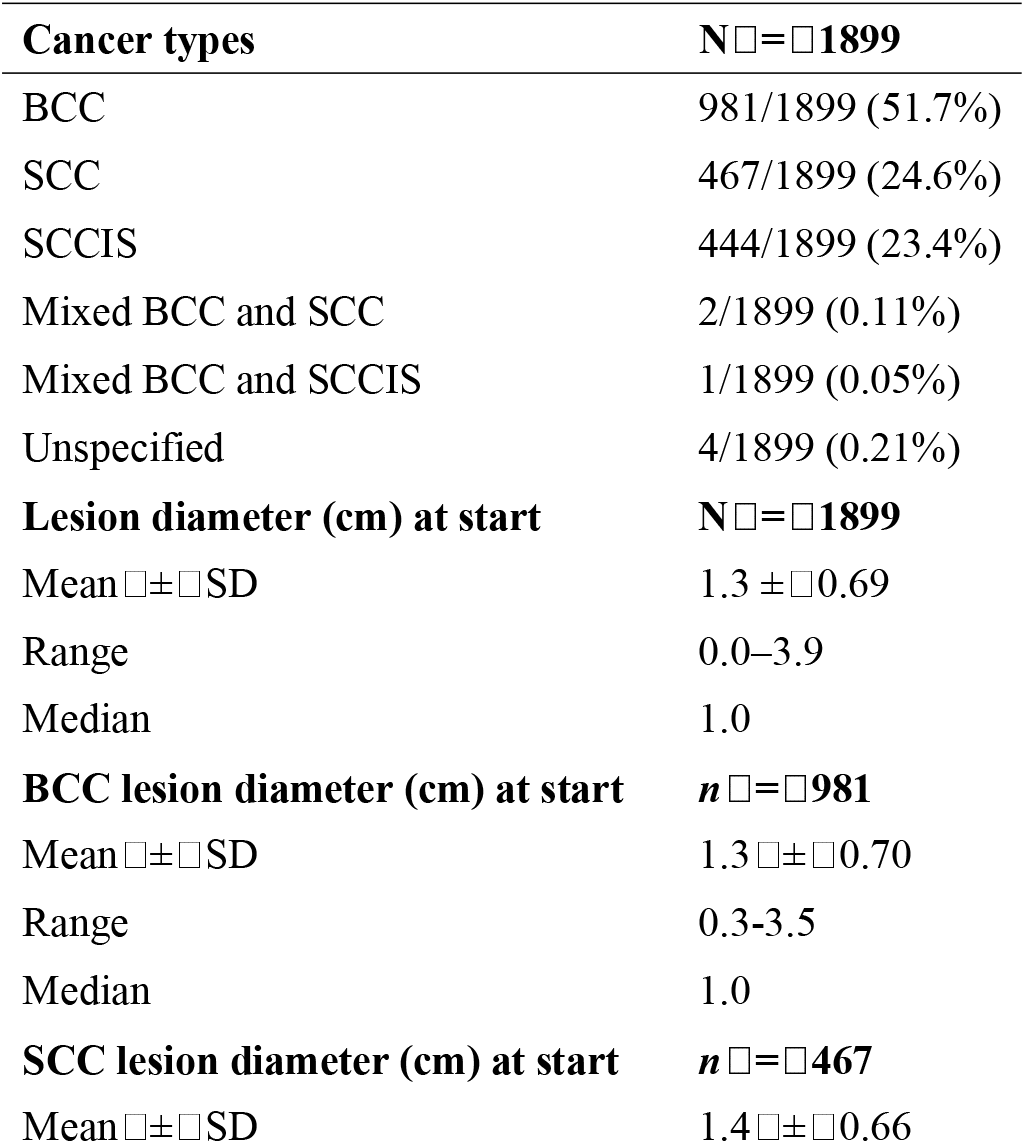
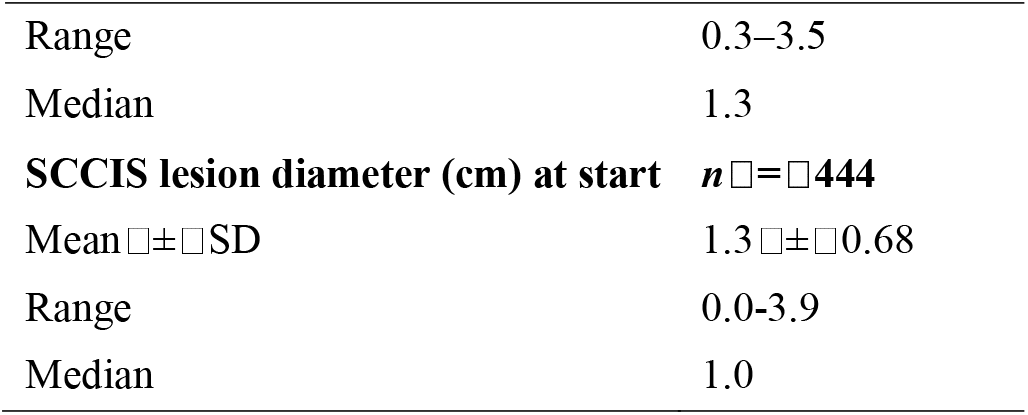
Cancer types and initial lesion size at initiation of treatment

**Table 6.** Diameter of NMSC lesions

| Diameter | BCC only | SCC only | SCCIS only | BCC & SCC | BCC & SCCIS | Total |
| --- | --- | --- | --- | --- | --- | --- |
| 0 to <2 cm | 766 | 342 | 338 | 1 | 1 | 1448 |
| 2 to <4 cm | 189 | 114 | 91 | 1 | 0 | 395 |
| Unspecified | 26 | 11 | 15 | 0 | 0 | 52 |
| **Total** | **981** | **467** | **444** | **2** | **1** | **1895<sup>a</sup>** |

<sup>a</sup>4 lesions had unspecified histology.

### 4.1 Local Control (LC)

Absolute LC was achieved in 99.7% of the lesions after an average of 7.5 weeks of treatment, with a stable control rate of 99.6% when the follow-up duration was over 12 months [Table 7]. Six lesions recurred at a mean of 6.83 months post treatment. Overall, Kaplan-Meier (KM) LC was 99.41% at the maximum follow-up time of 63.6 months (5 years) [Figure 1]. When comparing lesions by histology at maximum follow-up [Figure 2], KM LC for BCC and SCC were comparable at 99.24% and 99.16%, respectively. KM LC for SCCIS was 100% at maximum follow-up of 62.1 months. Log-rank comparison of KM LC between histologic subtypes (BCC, SCC, SCCIS) was not statistically significant (p= 0.2440, alpha= 0.05)

**Table 7.**
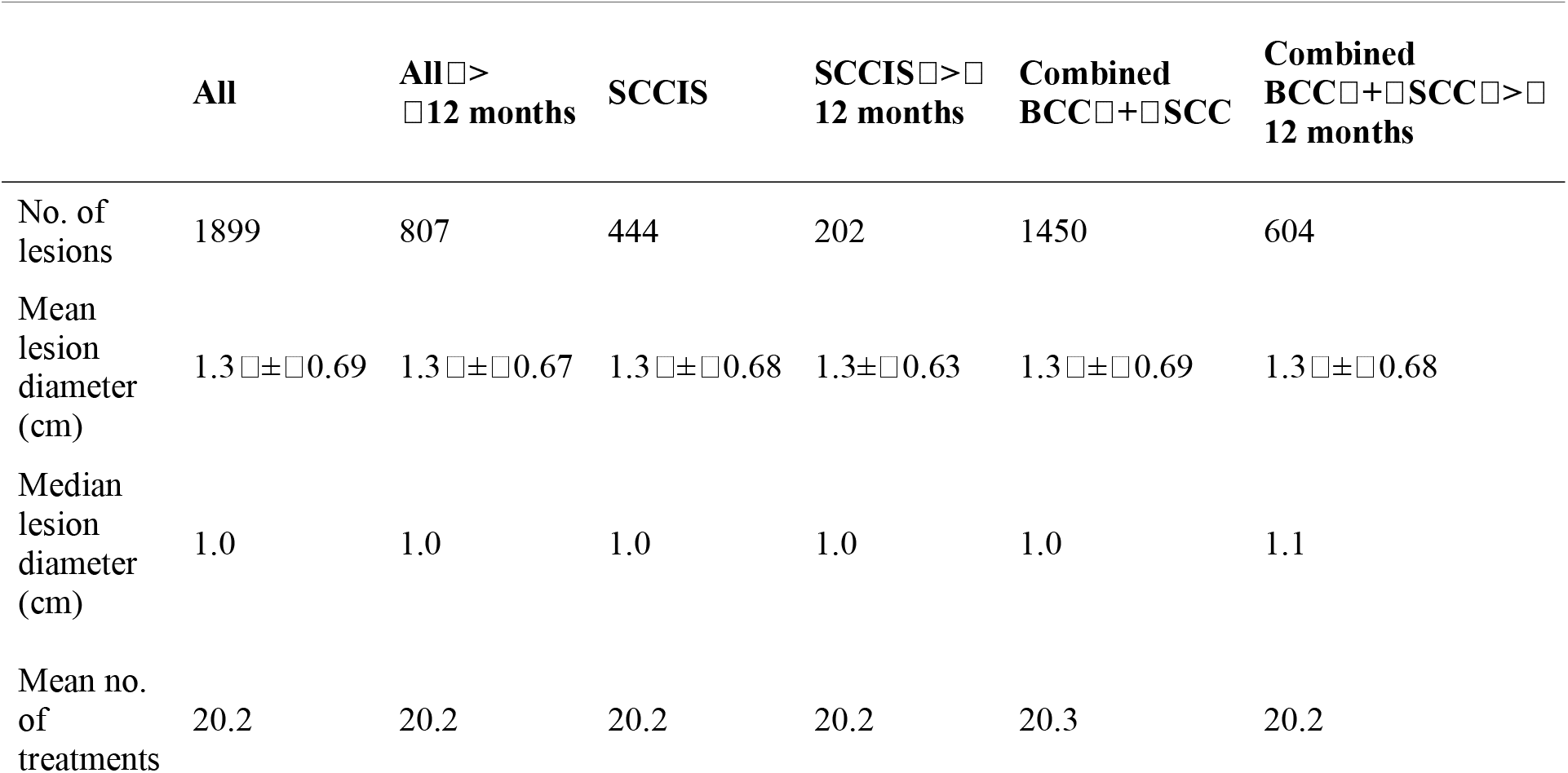
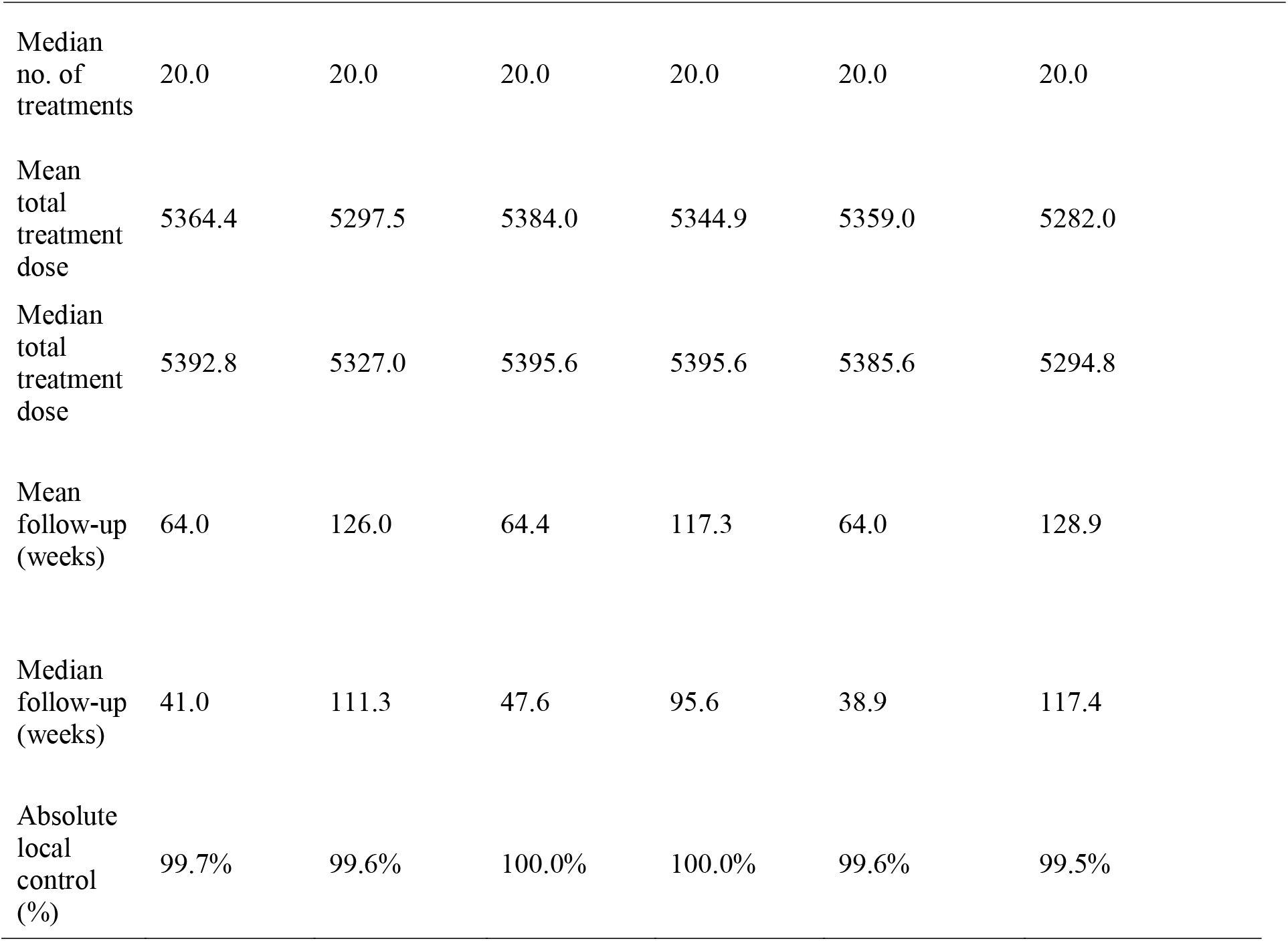
Control rates by tumor type and length of follow-up

**Figure 1:**
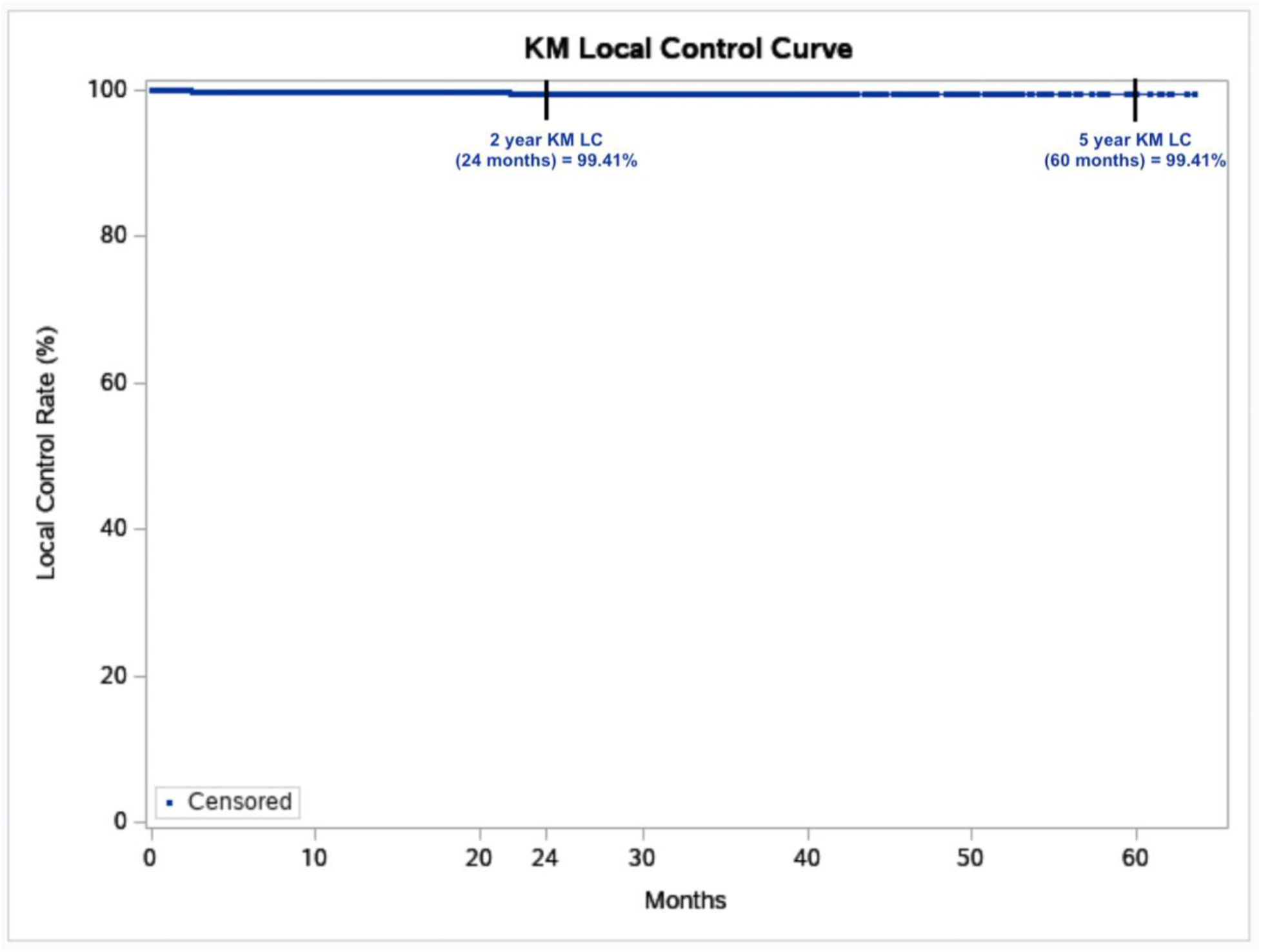
2 year and 5 year Kaplan-Meir (KM) local control (LC) for all 1899 lesions treated with Image-Guided Superficial Radiation Therapy from 2016 to 2022. Dots represent censored events.

**Figure 2:**
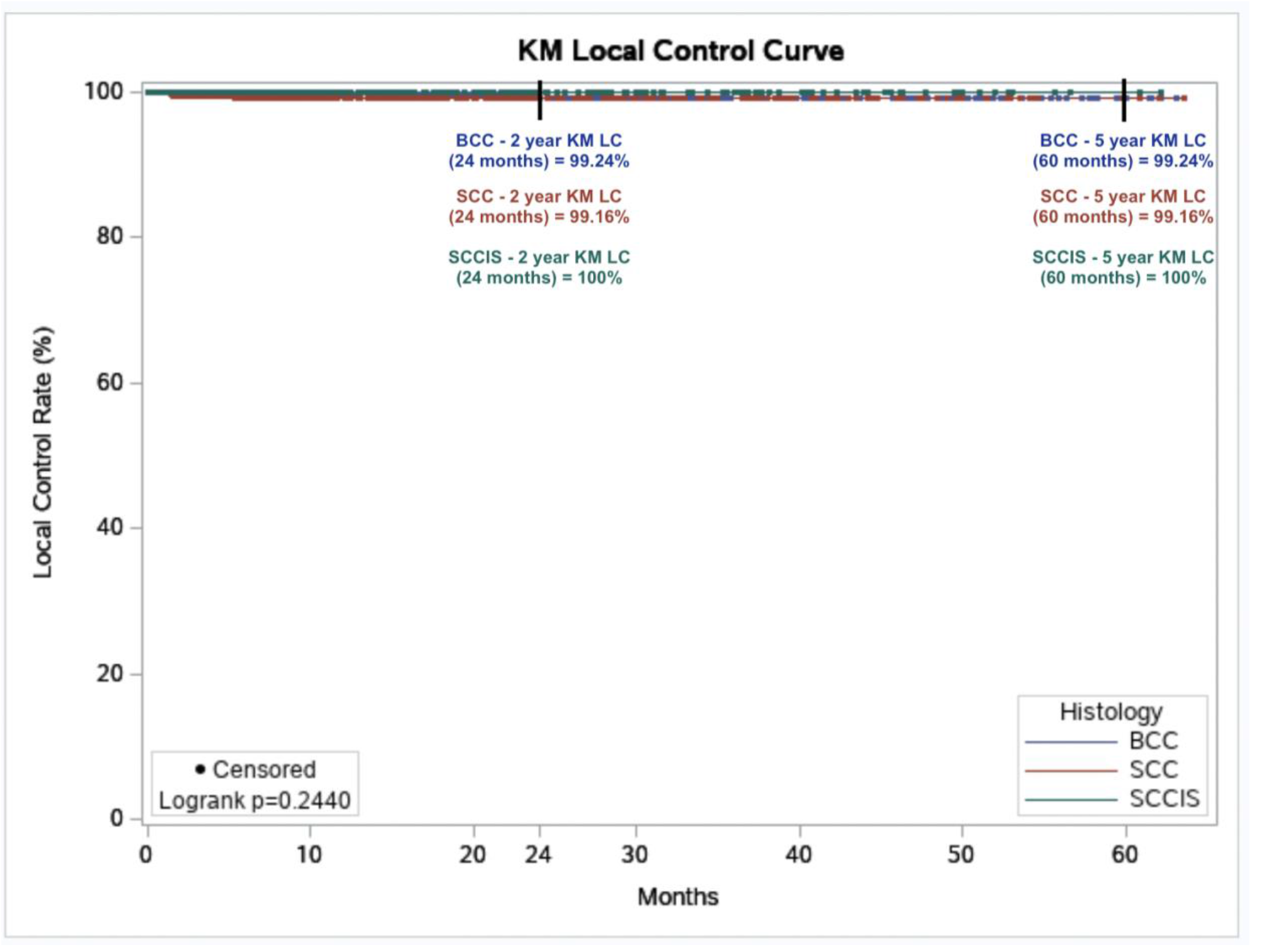
Kaplan-Meir (KM) local control (LC) by histology in all 1899 lesions treated with Image-Guided Superficial Radiation Therapy from 2016 to 2022.

### 4.2 Response

Among 1196 lesions with RTOG toxicity grade data available, 61 lesions (5.1%) were RTOG grade 3 or 4 in severity [Table 8]. 94.9% of lesions received RTOG grade of 1 or 2, consisting of mild or moderate self-resolving symptoms. The most common side effects displayed in patients was erythema, dryness followed by dry desquamation; however, some patients experienced ulceration and moist desquamation which did not affect lesion control. A median of 20 fractions was delivered at a mean total treatment dose of 5364.4 cGy with a mean follow-up of 65.5 weeks.

**Table 8.** Safety—by lesion based on RTOG criteria

| Characteristic | Grade | Description | (n = 1196)<sup>a</sup> |
| --- | --- | --- | --- |
| Highest RTOG toxicity grade | 1 | Follicular, faint, or dull erythema; epilation; dry desquamation; decreased sweating | 843/1196 (70.5%) |
|  | 2 | Tender or bright erythema, patchy moist desquamation; moderate edema | 292/1196 (24.4%) |
|  | 3 | Confluent, moist desquamation other than skin folds; pitting edema | 50/1196 (4.2%) |
|  | 4 | Ulceration, hemorrhage, necrosis | 11/1196 (0.9%) |

*RTOG* (Radiation Treatment Oncology Group)
<sup>a</sup>703 lesions missing *RTOG* grading

## 5.0 Discussion

The 2 year and 5 year absolute LC observed in this study are identical, which is consistent with the generally accepted premise that most failures in NMSC occurs within 2-3 years [27]. This study reinforces that the dosing regimen employed, along with the use of high-resolution ultrasound guidance, allow for achieving outstanding local control.

The most common and current curative radiation regimen in the hospital and cancer center setting consists of 180-200 cGy administered daily five times per week for 30-36 treatments to total cumulative doses of 5400-7200 cGy for all tumors [24]. For the skin, small fields may be treated with a hypofractionated regimen of 2-4 times weekly at higher fractions of 220-400 cGy [24]. In the aforementioned study, the protocol initially consisted of 255 cGy for 20 total fractions, with 50 kV given for lesion depth less than 1.5 mm and 70 kV for lesion depth greater than 1.5 mm three times a week [24]. The protocol has evolved over the years, in which a more detailed protocol that specified time dose fractionation (TDF) number/dose/fractionation was developed in 2019 based on ultrasound depth and tumor type. This protocol recommends a fractionation dose range of 245-279 cGy for 20 fractions 3-4 times a week to achieve a therapeutic biological dose range of 90-99 or greater TDF number using 50, 70, or 100 kV energy. Higher doses per fraction and/or more fractions were recommended for larger, deeper, and high-risk lesions [24]. Nonetheless, energy administered is contingent upon anatomic location, histology, lesion depth, and skin curvature. This study predominantly uses the updated protocol and achieves comparable excellent results to the Yu study. Additionally, these results compare favorably with MMS. Our study affirms its efficacy and safety as our results showed a 99.7% control rate, which is comparable to the 99.3% control rate found in the multi-institutional SRT study led by Yu [24] and patients experienced similar self-limiting side effects with a majority of patients receiving a RTOG score of 1.

The acute toxicity in this study showing <1% Grade 4 RTOG toxicity is also compatible with results reported by Yu et al [24]. The approximately 4% Grade 3 RTOG toxicity, however, while still low, may imply there may be slightly more toxicity, but is still very much within safe parameters with the use of a more modern 2019 protocol detailed above. A possible explanation for the increase in Grade 3 RTOG toxicity over that reported by Yu et al [24] is the slightly increased median dose from 5188 cGy to 5393 cGy, which also seems to imply an increase in local control from 99.3% to 99.7%. Although this improvement is <1%, the sheer numbers of annual lesions with NMSC in the United States alone (approximately 5.3 million causes) theoretically translates to ∼50,000 more lesions controlled. The lesions of patients treated from 2019 onward using the updated protocol in the original 2021 Yu study was 30% (876/2917) (internal data courtesy of Dr. Yu). Whereas in this study, roughly 74% (1398/1899) of the lesions were treated after 2019. IGSRT has a high safety profile with limited, self-resolving side effects such as minimal pain, swelling, desquamation, and radiation dermatitis, making it a safe, cost-effective option for NMSC treatment. Given the overall trend towards less/non-invasive and non-surgical options in medicine, IGSRT remains a promising alternative to MMS and other surgical options and should be considered more readily for patients with early-stage NMSC, advanced age, and patients with medical comorbidities, e.g., diabetes, stasis dermatitis, chronic edema that may render them poor surgical candidates/have poor wound healing capability. Additionally, there are implications for improved care through improved quality of life via prevention of disfigurement (as well as associated pain and pruritus), especially in scar/keloid individuals, e.g., darker pigmented individuals with substantial evidence for its effectiveness in treating recurrent/resistant keloid scars [25]. It also has the potential to address disparities to access to dermatologic care as the patient may receive IGSRT from other qualified personnel, e.g., Board-Certified radiation therapists and IGSRT is often covered by insurance. This option may be especially beneficial in underserved areas, i.e., rural areas or areas underserved by dermatologists. Additionally, this non-invasive treatment option may be advantageous for those with scar/keloid-prone skin [25], such as those with darker pigmented skin, e.g., African Americans, Asians, and Hispanics. Currently, high-precision radiation oncology aims to optimize tumor coverage without sacrificing normal tissues. IGSRT with ultrasound assistance is one example of personalized oncology whereby precise high-dose delivery can be achieved at the desired superficial level.

## 6.0 Conclusion

The use of image guidance with high resolution dermal ultrasound has advanced superficial radiation therapy as it enables more precise depth coverage, greater local control/cure rates, lower complication rates, and better cosmesis/function. IGSRT should be considered first-line for treating early-stage NMSC tumors as cure rates have been shown to be effective in all NMSC on early follow-up and has the potential to be superior (with more follow-up data) to traditional SRT and surgery [24]. IGSRT may be useful in improving disparities in underserved communities, persons of color, those of low socioeconomic status and underprivileged segments of the population.

## Data Availability

All data generated or analyzed during this study are included in this published article [and its supplementary information files].

## Statements and Declarations

## Ethics Approval

The study protocol was reviewed by an IRB committee (WIRB-Copernicus Group) and determined to be exempt from IRB approval under 45 CFR 46.104 (d)(4) and was performed in compliance with the pertinent sections of the Helsinki Declaration and its amendments.

## Consent

N/A

## Author Contributions

The authors whose names appear on the submission have contributed sufficiently to the manuscript and approved the final submitted version of the manuscript.

